# Is Second dose of vaccination useful in previously SARS-CoV-2 infected Health Care Workers?

**DOI:** 10.1101/2021.06.22.21257103

**Authors:** Gauthier Pean De Ponfilly, Benoît Pilmis, Iheb El Kaibi, Nathalie Castreau, Sophie Laplanche, Alban Le Monnier

## Abstract

**Summary:** Vaccines are the most important public health measure to protect people from COVID-19 worldwide. In addition, healthcare workers account for a large number of infected people. Then, protecting this population from COVID-19 seems crucial in the preservation of healthcare systems. In a context of few doses available, serological assays could be useful to decide whether one or two doses are needed. Our results show that a first dose of BNT162b2 mRNA vaccine seems to act as a boost after SARS-CoV-2 infection in healthcare workers with a previous SARS-CoV-2 infection and a second dose might not be required.

## Introduction

Since the early 2020, a large pandemic due to the new and emergent coronavirus (SARS-CoV-2) spreads all over the world. While some countries have been able to contain the pandemic with a combination of public health measures (extensive testing followed by isolation and tracing of contacts, near-universal masking, and target quarantines), most countries have failed to stop the pandemic. Much hope now resides in the potential of SARS-CoV-2 vaccines to reduce the risk of disease and infection. Strategies aimed to maximize the early impact of vaccination in a context of few doses available [1]. As soon as early January 2021, large vaccination campaign started in France first for populations at risk of severe COVID-19 but rapidly also for Healthcare workers (HCWs) more than 50 years or with underlying diseased, then for all voluntary HCWs. Indeed, numerous studies have underlined the risk of nosocomial transmission to and from healthcare workers in hospital settings [2,3].

Then, the issue of vaccination of patients and HCWs with a history of recent or late SARS-CoV-2 related infection has become a major issue to reduce this risk. Currently, French guidelines recommends vaccination in those cohort that are recovered from COVID-19 since six months and to offer them only one dose [4]. Moreover, serological assays are not included in the decision-making strategy of vaccination.

We aimed to provide data on immune response induced by SARS-CoV-2 vaccines in real world context that can support this choice. Here, we reported kinetics of production of antibodies directed to the Spike protein of SARS CoV-2 after first, then second vaccine dose (conventional prime-boost strategy recommended) in Health Care Workers.

## Material and methods

All HCWs currently working at the Groupe Hospitalier Paris Saint-Joseph (Paris, France) and eligible to vaccination were proposed to be included in an observational cohort study after first then second dose of the BNT162b2 mRNA COVID-19 vaccine (Pfizer / BioNTech, Mainz, Germany).

They were divided in two groups: HCWs with a previous history of SARS-Cov-2 infection (COVID-19 (+)) and HCWs with no history of SARS-Cov-2 infection (COVID-19 (-)) according to the result of RT-PCR and/or serology to confirm previous contact with SARS-CoV-2. Minimal information were collected like sex, age, dates of first and second vaccine doses.

After agreement, HCWs have first serology 28 days after the first dose before receiving the second dose, then a second serology 21 to 28 days after.

HCWs with a history of COVID-19 came from the ongoing PERSOCOVID longitudinal observational cohort study. Briefly, these colleagues have benefited of a longitudinal serological follow-up since the onset of the disease. For each of them, the level of antibodies directed to Spike protein before vaccination was known. No COVID-19 recently diagnosed in the last three months were included in the study, according to French health agencies recommendations.

Serological assays used the quantitative test (SARS-CoV-2 IgG II Quant, Architect System, Abbott) for detecting serum antibodies directed to receptor-binding domain (RBD) of the Spike-1 protein of SARS-CoV-2. Quantitative results are converted in AU/mL. Dilution was performed for each result beyond 40,000 AU/mL according to the manufacturer’s instructions. The positivity threshold was 50 AU/mL according to manufacturer’s instructions.

For the statistical comparison, a Shapiro-Wilk test for normality of distribution was first performed and a t-student test was performed for continuous variables, using the R software version 3.1.3[4]. Significance was considered if p<0.05.

## Results

Seventy-three HCWs gave consent to participated to this study and distributed in 29 (39.7%) COVID-19 (+) and 44 (60.3%) COVID-19 (-). No case of severe COVID-19 was recruited in this study and median [IQR] age of the included HCWs was 43.5 [34 – 56.5] years old. The median time from first dose to serology was 25 days and 28 days from second dose to second serology. We first reported a significant difference in anti-Spike antibody levels (median [IQR]) around 46-fold higher in COVID-19 (+) (28111 [15032-33967] AU/mL) than in COVID-19 (-) (642 [338-1170] AU/mL) (*p* < 0.001) after the first dose. Antibody levels produced after the first dose in COVID-19 (-) are quite similar than those observed in COVID-19 (+) (324 [116-630] AU/mL) not yet vaccinated. In the latter, the antibody levels increased on a mean by 88-fold after the first vaccine dose.

After the first vaccine dose, COVID-19 (-) presented a median level of IgG anti-Spike at 617 [338-1147] AU/mL and their median level increases significantly 28 days after their second dose at 9711 [4978-15473] AU/mL (*p* < 0.001). Previous infection seemed to be analogous to natural immune priming.

After the second dose, COVID-19 (-) significantly increased their antibody levels (10491 [5341-16193] AU/mL) without, however, reaching the levels generally observed for COVID-19 (+) after a single dose. For COVID-19 (+), we observed that the second dose did not significantly increase their antibody level (35459 [11565-38500] AU/mL; *p*=0.18).

## Discussion

Following a single dose of BNT162b2, HCWs with a previous SARS-CoV-2 infection have significantly higher antibody response than naive HCWs. The first dose of the vaccine seems to act as a boost after SARS-CoV-2 infection and a second dose might not be required in case of previous SARS-CoV-2 infections.

A delayed booster beyond the 30 days initially recommended for the BNT162b2/Pfizer vaccine could then be proposed. This is an important element in the current context of few doses available to accelerate vaccine rollout [1].

Furthermore, COVID-19 is associated with an increase in HCWs absenteeism making the management of a public health crisis even more complicated [5].

However, we also reported a paradoxically lower response than those expected for one COVID-19 (+). This HCW presented a positive RT-PCR and a positive serological assay confirming previous contact with SARS-CoV-2. This suggests that the use of serological tests could be useful to confirm adequate vaccine response without, however, the correlate of protection value. Indeed, here no neutralization assays have been carried out because they are difficult to set up on a routine basis [6]. For HCWs with a previous SARS-CoV-2 infection, a determination of the antibodies level produced after the first dose can be proposed just before the second dose to decide whether or not it is necessary.

Kinetics of natural anti-Spike antibody levels decrease more or less rapidly according to initial level of IgG produced. In some cases, anti-Spike antibodies become undetectable beyond six or eight months for some HCWS or some patients [7,8]. This also raises the question of the durability of the antibodies produced either after prime or boosted vaccination as for COVID-19 course and the impact of high initial antibodies levels.

Our study provides additional information on the positioning and usefulness of serology in the COVID-19 vaccine strategy in order to maximize coverage and impact.

## Data Availability

All data are available

## Acknowledgement

We thank all colleagues who accept to participate to this study, in particular staff of the Occupational Medicine Department: Morgane PAUILLAT, Mathilde HERR, Dr Priscilla SAVIN, and Dr Patricia MALARDY.

---

**Figure 1.**
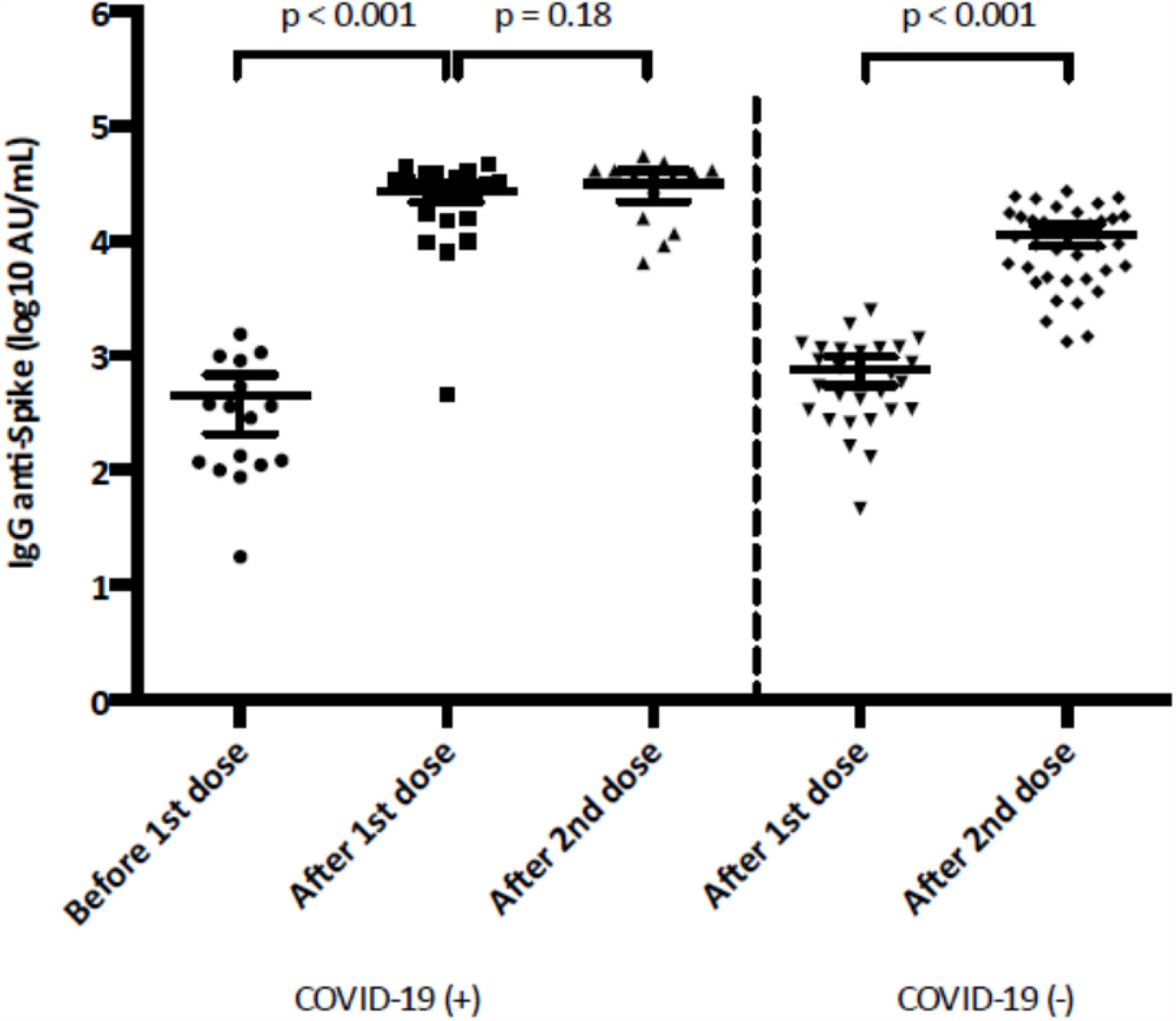
Evolution of anti-Spike IgG titer expressed in log10 AU/mL during the vaccination protocol, according to history of COVID-19.

